# MCNET: Multi-Omics Integration for Gene Regulatory Network Inference from scRNA-seq

**DOI:** 10.1101/2023.05.29.23290691

**Authors:** Ansh Tiwari, Sachin Trankatwar

## Abstract

Deep learning has emerged as a powerful approach in various domains, including biological network analysis. This paper investigates the advancements in computational techniques for inferring gene regulatory networks (GRNs) and introduces MCNET, a state-of-the-art deep learning algorithm. MCNET integrates multi-omics data to infer GRNs and extract biologically significant representations from single-cell RNA sequencing (scRNA-seq) data. By incorporating attention mechanisms and graph convolutional networks, MCNET captures intricate regulatory relationships among genes. Extensive benchmarking on diverse scRNA-seq datasets demonstrates MCNET’s superiority over existing methods in GRN inference, scRNA-seq data visualization, clustering, and simulation. Notably, MCNET accurately predicts gene regulations on cell-type marker genes in the mouse cortex, validated by epigenetic data. The introduction of MCNET paves the way for advanced analysis of scRNA-seq data and provides a powerful tool for inferring GRNs in a multi-omics context. Moreover, this paper addresses the integration of multiomics data in gene regulatory network inference, proposing MCNET as a method that efficiently analyzes and visualizes homogeneous gene regulatory networks derived from diverse omics data. The inference capability of MCNET is evaluated through extensive experiments with simulation data and applied to analyze the biological network of psychiatric disorders using human brain data.

## 1. Introduction

Understanding many biological processes requires knowledge not only about the biological entities themselves but also the relationships among them. For example, processes such as cell differentiation depend not only on which proteins are present, but also on which proteins bind together. A natural way to represent such processes is as a graph, also called a network, since a graph can model both entities as well as their interactions. Recent advances in experimental high-throughput technology have vastly increased the data output from interaction screens at a lower cost and resulted in a large amount of such biological network data (Reuter et al., 2015). The availability of this data makes it possible to use biological network analysis to tackle many exciting challenges in bioinformatics, such as predicting the function of a new protein based on its structure or anticipating how a new drug will interact with biological pathways. This wealth of new data, combined with the recent advances in computing technology that has enabled the fast processing of such data (Goodfellow et al., 2016), has reignited interest in neural networks (Sokolov et al., 2015; Parker, 1985; LeCun, 1985; Rumelhart et al., 1986) which date back to the 1970s and 1980s, and set the stage for the emergence of deep neural networks, a.k.a deep learning, as a new way to address these unsolved problems.

Biological systems on different levels of organization, from organelles and single cells to tissues, organs and entire organisms, constantly sense the environment and modulate their behavior to ensure optimal performance and fitness (López-Barneo et al., 2001; Rolland et al., 2006; Veal et al., 2007). The sensing of the environment is accomplished via numerous molecular mechanisms which ultimately result in coordinate activation and suppression of, often multiple, regulatory cascades affecting different and mutually dependent cellular processes. By propagating the perceived signal, the expression levels of genes coding for transcription factors (TFs) are adequately altered, leading to changes in the levels of transcripts encoding enzymatic proteins which affect metabolism and organism’s tasks (Jacob & Monod, 1961).

Therefore, accurate reconstruction of the complete set of regulatory interactions, forming gene regulatory networks, is one of the key tasks in systems biology (Karlebach & Shamir, 2008). Gene regulatory networks have an important role in every process of life, including cell differentiation, metabolism, the cell cycle and signal transduction. By understanding the dynamics of these networks we can shed light on the mechanisms of diseases that occur when these cellular processes are dysregulated. Accurate prediction of the behaviour of regulatory networks will also speed up biotechnological projects, as such predictions are quicker and cheaper than lab experiments Karlebach & Shamir (2008).

In biological systems, the gene regulatory interactions are transitory, as they depend on different factors, including: developmental, environmental, as well as internal, given by the genetic make-up of the organism. the expression of cognate genes is integrated in layers of iterative regulatory networks that ensure the performance not only of the whole cell, but also of the bacterial population, and even of the entire microbial community, in a changing environment (Cases & de Lorenzo, 2005).

High-throughput technologies for simultaneous measurement of gene expression have been used to capture the transitory behavior of thousands of genes upon internal and external perturbation in different biological systems, from bacteria and yeast to algae, plants and animals (Schulze & Downward, 2001; Blencowe et al., 2009; Rehrauer et al., 2009). The gathered gene expression levels reflect the underlying regulatory relationships, and, thus, can readily be used to reconstruct the operational regulatory networks.

With the increasing number of performed time-series experiments methods are needed to extract gene regulatory networks supported by all gathered data sets simultaneously. These experiments are over different time domains, with different sampling frequency under various conditions and conducted in different laboratories, which may affect the success of network reconstruction (Sima et al., 2009). In addition, each of these experiments is usually accompanied by a corresponding reference control experiment, whose profiles are used to determine differential gene behaviors (García de la Nava et al., 2004; Rapaport et al., 2013).

Reconstruction of gene regulatory networks is a classical problem in computational systems biology and various methods based on different sets of assumptions and applicable on data from particular experiments have been proposed, critically assessed and systematically reviewed (Hempel et al., 2011; Marbach et al., 2012; Omony, 2014).

In general, inference of gene regulatory networks begins with application of a similarity measure of choice on the investigated data set, resulting in a square similarity matrix. This similarity matrix can be sparsified by retaining only the values which are statistically significant after multiple hypothesis testing. A number of computational models (Rumelhart et al., 1986; Huynh-Thu et al., 2010; Chan et al., 2017; Matsumoto et al., 2017; Papili Gao et al., 2018) have attempted to incorporate GRN inference into their single-cell data analysis models. Current methods solely based on single-cell RNA sequencing (scRNA-seq) data also have explicit limitations. For example, it is common for GRN inference algorithms to use statistics algorithms that focus on the co-expression networks instead of decoding the casual relationships among TFs and their corresponding target genes (Chan et al., 2017; S. Kim, 2015). A number of computational models (Rumelhart et al., 1986; Huynh-Thu et al., 2010; Chan et al., 2017; Matsumoto et al., 2017; Papili Gao et al., 2018; Moerman et al., 2019) have attempted to incorporate GRN inference into their single-cell data analysis models.(Matsumoto et al., 2017; Papili Gao et al., 2018) or tree-based models (Huynh-Thu et al., 2010; Moerman et al., 2019) and it is generally hard to directly generalize these approaches to more comprehensive nonlinear frameworks and benefit from the computational power that the deep learning model brought to us. One class of these methods relies on side measurements such as single-cell chromatin accessibility or transcription factor (TF) binding motifs (Kamimoto et al., 2023). However, these measurements often require more complicated experimental designs and could also introduce additional noise as these data could come from different experiments.

To address the above problems, we present MCNET, a deep generative model that can jointly embed the gene expression data and simultaneously construct a GRN that reflects the inner structure of gene interactions in single cells. To implement such an idea, we heavily based our work on the work of [29], which generalized a popular approach, called the structural equation model (SEM), that infers the causality using a linear model, and implemented the exact technique that was desired for our project. (Shu et al., 2021) hypothesised that by adding proper mathematical constraints, part of the neural network architecture could be used to predict the GRN of the scRNA-seq data. A previous study by (Lin et al., 2017) showed that more accurate cell representations could be achieved by guiding the neural network architecture with a GRN structure derived from the literature and databases. In this Article, we show that the neural network architecture can reflect GRN structure by properly designing the neural network layer with a reliance on multi omic data. Integration of multi-omic datasets in a Structural Equation Modelling neural network was based on the work of (Picard et al., 2021). The neural network architecture can be inferred jointly with the training of the weights of the neural network in an end-to-end manner.

We evaluate the performance of MCNET for various single-cell tasks such as GRN inference, scRNA-seq data visualization, cell-type identification and cell simulations on several benchmark datasets. We first show that MCNET is able to achieve better performance on the GRN inference task compared with the state-of-the-art algorithms on several popular benchmark datasets. We also apply MCNET to another single-cell dataset without the ground-truth GRN measured, and provide extensive evidence extracted from the single-cell DNA methylation and open chromatin data to demonstrate the accuracy and efficiency of our algorithm. Moreover, we also evaluate the quality of the single-cell representation regularized by the GRN structure. We find that MCNET can achieve comparable or better performance compared with current state-of-the-art methods on the tasks of visualization and cell-type identification on various benchmark datasets.

## 2. Integration of Multi Omic data

The advent of powerful and inexpensive screening technologies (Misra et al., 2019) recently produced huge amounts of biological data that opened the way to a new era of therapeutics and personalized medicine (Ahmed, 2020) Treatment efficiency and adverse effects can differ vastly between individuals due to differences in age, sex, genetics and environmental factors (e.g., anthropometric and metabolic statuses; dietary and lifestyle habits (Burney & Lakhtakia, 2017; Jaccard et al., 2017) The aim of precision medicine is thus to design the most appropriate intervention based on the biological information of each individual (Tebani et al., 2016). Clinical information and omics data can be directly retrieved from databases or collected with screening technologies for disease (Menyhárt & Győrffy, 2021), class prediction (Hasin et al., 2017), biomarkers discovery (Sun & Hu, 2016), disease subtyping (Menyhárt & Győrffy, 2021), improved system biology knowledge (Dahal et al., 2020), drug repurposing and so on. Each type of omics data is specific to a single “layer” of biological information such as genomics, epigenomics, transcriptomics, proteomics, metabolomics, and provides a complementary medical perspective of a biological system or an individual (Misra et al., 2019). In the past, single-omics studies were done in hope of discovering the causes of pathologies and helping select an appropriate treatment. We now realize that such approaches are overly simplistic. Most diseases affect complex molecular pathways where different biological layers interact with each other. Hence the need for multiomics studies that can encompass several layers at once and draw a more complete picture of a given phenotype (Tian et al., 2014) With multiple omics, faint patterns in gene expression data can be reinforced with epigenomics (Zarayeneh et al., 2017) for example. Complementary information can be exploited to better explain classification results (Rappoport et al., 2020), improve prediction performances (Sharifi-Noghabi et al., 2019), (Tini et al., 2017) or understand complex molecular pathways (Akhmedov et al., 2017) that would be out of grasp for single-omics studies. However, multi-omics studies include data that differ in type, scale and distribution, with often thousands of variables and only few samples. Additionally, biological datasets are complex, noisy, with potential errors due to measurement mistakes or unique biological deviations. Discovering pertinent information and integrating the omics into a meaningful model is therefore difficult and a great number of methods and strategies have been developed in recent years to tackle this challenge (Menyhárt & Győrffy, 2021), (Higdon et al., 2015). If the integration is not done correctly, adding more omics might not result in a significant increase of performance, but will increase the complexity of the problem along with computational time.

### 2.1. Main integration strategies

From multiple omics datasets, each having the same rows representing samples (patients, cells) and different columns representing biological variables grouped by omics (gene expression, copy number variation, miRNA expression, etc.), different goals could be achieved such as sample classification, disease subtyping, biomarker discovery, etc. Machine learning (ML) models are commonly used to analyze complex data, but the integration of multiple noisy and highly dimensional datasets is not straightforward. Hence, multiple integration strategies have been developed, each one of them having pros and cons. Assuming each dataset has been pre-processed according to its omics data, the datasets could simply be assembled with sample wise concatenation and the resulting matrix used as input to ML models. But in practice, most ML models will struggle to learn on such a complex dataset, particularly if the number of samples is low. Other strategies rely on transforming or mapping the datasets to reduce their complexity, either independently or jointly. An opposite strategy can also be adopted, which does not combine data and analyzes each omics dataset separately. The prediction of each model is assembled afterward for a final decision. Finally, the hierarchical strategy integrates the omics datasets by taking into account the known regulatory relationships between omics as presented by the central dogma of molecular biology (CRICK, 1970).

### 2.2. Hierarchical integration

A challenge in system biology is to understand the modular organization structured at the molecular level. A new trend is to incorporate these regulatory effects in the integration strategy to better reflect the nature of multidimensional data. Hierarchical strategy bases the multi-omics integration on the inclusion of the prior knowledge of regulatory relationships between the different layers. For example, a strategy for genotype-phenotype integration based on existing knowledge of cellular subsystems could follow this logic: genotypic variations in nucleotides can give rise to change in gene expression or functional changes in proteins which in turn could ultimately affect the phenotype. Therefore, hierarchical integration strategies often use external information from interaction databases and scientific literature. Moreover, because omics are organized in sequential fashion, the challenges of multi-omics integration are not exacerbated and can be dealt with separately for each dataset. Some methods for supervised hierarchical integration include Bayesian analysis of genomics data (iBAG) (Wang et al., 2013), linear regulatory modules (LRMs) (Zhu et al., 2016), and Assisted Robust Marker Identification (ARMI) (Chai et al., 2017), and Robust Network (Wu et al., 2018). Hierarchical integration methods are often designed to study specific regulatory relationships. For example, iBAG has been developed to investigate associations between epigenetic and gene expression regulation. The framework uses hierarchical modeling to combine the data from methylation and gene expression to study the associations with patient survival. Robust Network has developed an approach for modeling the gene expression (GE) and copy number variation (CNV) regulation that describe the dominant cis-acting CNV effects compared to trans-acting CNVs. This approach could be extended to other regulation relationships such as gene expression by methylation and microRNAs. Additionally, hierarchical integration can be used to infer gene regulatory networks (GRN) from multi-omics datasets.

## 3. Methods

### 3.1. Notations

We describe the notations used throughout this paper. Let *G* denote a matrix of gene expression, *G* ∈ ℝ^*N×P*^, where *N* is the number of samples and *P* is the number of genes in microarray data. The vector of gene expression at the *i*th gene is denoted as *g*_*i*_ ∈ ℝ^*N*^, and *G*_−*i*_ represents the matrix that contains gene expressions other than gene *i, G*_−*i*_ ∈ ℝ^*N×*(*P* −1)^. The matrices of CNV and DNA methylation data are denoted as *C* ∈ ℝ^*N×V*^ and *D* ∈ ℝ^*N×M*^, respectively. We suppose that CNVs or DNA methylations can be annotated to nearby genes (upstream or downstream of the gene) in the same chromosome. The gene annotations of the CNV and DNA methylation to gene *i* are represented as the matrices 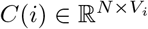 and 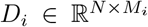, respectively, where *V*_*i*_ and *M*_*i*_ are the numbers of CNVs and DNA methylations that are annotated to gene *i*.

The regulatory relationships between genes are represented by an adjacency matrix of gene expression *B* ∈ ℝ^*P ×P*^, and integrative interactions of multi-omics data other than gene expression are expressed by their own biadjacency matrices. In MCNET, the interactions of CNVs and DNA methylations to genes are described as *BC* ∈ ℝ^*V ×P*^ and *BD* ∈ ℝ^*M×P*^, respectively. We assume that there is no self-regulation in the gene regulatory network, i.e., *B*_*ii*_ = 0, where *i* = {1, …, *P* }.

### 3.2. Integrative gene regulatory network inference

We propose an Integrative Gene Regulatory Network inference (MCNET) method that infers a gene regulatory network from multi-omics data. The current state-of-the-art methods for integrative gene regulatory network inference using multiomics data, such as SGRN (Cai et al., 2013) and DCGRN (D.-C. Kim et al., 2014a), consider all the types of data as nodes in networks. In other words, nodes can indicate genes, CNVs, or DNA methylations. In contrast, MCNET constructs homogeneous gene regulatory networks where nodes represent only genes, which consequently makes it possible to apply most graph algorithms for further analysis. Figure 1 shows a simple integrative gene regulatory network, where a gene (G1) regulates another gene (G4) with biological processes of a CNV (CNV2) and a DNA methylation (DM2). The proposed method, MCNET, represents integrative gene regulatory networks with multi-layered adjacency matrices of the multiomics data. It constructs the adjacency matrix of gene expression and the biadjacency matrices of CNV and DNA methylation. The adjacency matrix of gene expression defines the basic structure of the transcriptional biological networks, and the biadjacency matrices of CNV and DNA methylation describe their integrative interactions on the gene regulations. For formulating the integration of the heterogeneous data into a standardised format, MCNET takes into account the interaction effects of CNVs and DNA methylations with genes. The integrative interactions between a gene i and its nearby CNVs and DNA methylations can be described by Fisher’s interaction model:

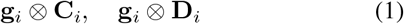

where ⊗ is an element-by-element multiplication. It explains different gene expression levels on the variations of CNVs or DNA methylations. Thus, the expression of gene *i* can be represented by a sparse linear model by incorporating not only other genes but also interaction effects of its nearby CNVs and DNA methylations. The gene expression (*g*_*i*_) for gene *i* is formulated by:

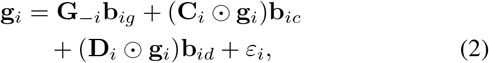

where *b*_*gi*_, *b*_*ci*_, and *b*_*di*_ are the coefficients of gene expressions other than gene *i*, CNVs, and DNA methylations of gene *i*, respectively. | *·* | is the L-1 norm, and the residual is denoted as *ε*_*i*_. The adjacency matrix *B* of the gene regulatory network is comprised of *b*_*gi*_ (1 ≤ *i* ≤ *P*) in (2), i.e., *B* = {*b*_*g*1_, …, *b*_*gP*_ }^⊥^.

**Figure 1.**
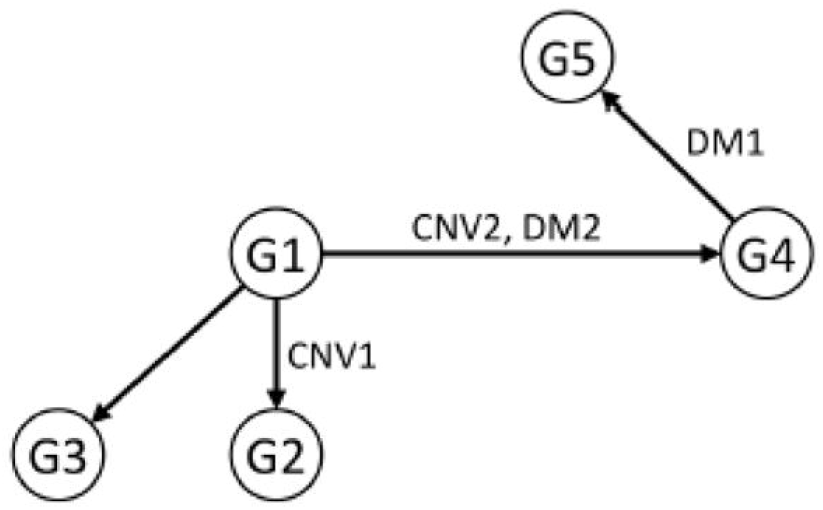
A simple integrative gene regulatory network. The interaction effects of copy number variation (CNV) and DNA methylation (DM) are incorporated in the gene regulatory network model

The biadjacency matrices of CNVs *BC* and DNA methylations *BD* are also constructed by *b*_*ci*_ and *b*_*di*_.

The integrative gene regulatory network can be inferred by optimizing the parameters of (2). The learning function *F* (*b*_*gi*_, *b*_*ci*_, *b*_*di*_) for the optimal parameters is obtained by using least squares with the sparse setting:

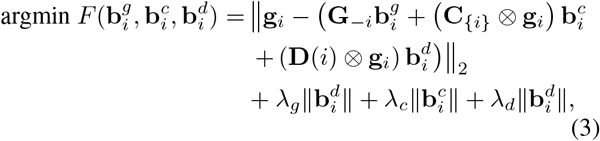

where *λ*_*g*_, *λ*_*c*_, and *λ*_*d*_ are hyper-parameters for sparsity regularization, and |*·*|_2_ is the L-2 norm. The optimization function can be considered as the following LASSO problem:

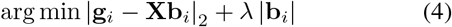

where *X* is the augmented matrix, *X* = {*G*_−*i*_, *C*(*i*)⊗*g*_*i*_, *D*_*i*_⊗ *g*_*i*_}. However, the number of genes in (*P* − 1) is much larger than the number of CNVs (*V*_*i*_) and DNA methylations (*M*_*i*_) associated with gene *i*. For instance, there are only a couple of CNVs (*C*_*i*_) or DNA methylations (*D*_*i*_) for a gene in the psychiatric disorder data that we used for the experiment in the paper, whereas the number of genes in *G*_−*i*_ is in the hundreds even after pre-processing. Thus, the solution of LASSO may tend to ignore most CNVs and DNA methylations despite their importance. Therefore, we solve the optimization problem in a stepwise manner. First, we identify significant genes that interact with gene *i* from *G*_−*i*_ by LASSO:

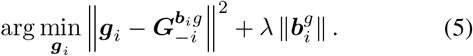

The matrix of 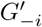 is constructed with the genes with non-zero coefficients. Secondly, we compute p-values of the variables in the following linear regression:

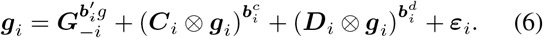

The coefficients of the genes, CNVs, and DNA methylations with p-values ≥ 0.05 are set to zeros. Then, the coefficients for genes are assigned to the adjacency matrix, and *b*_*ci*_ and *b*_*di*_ are assigned to the biadjacency matrices of *BC* and *BD* respectively. The procedure is described in Algorithm 1.

#### Algorithm 1 Algorithm 1

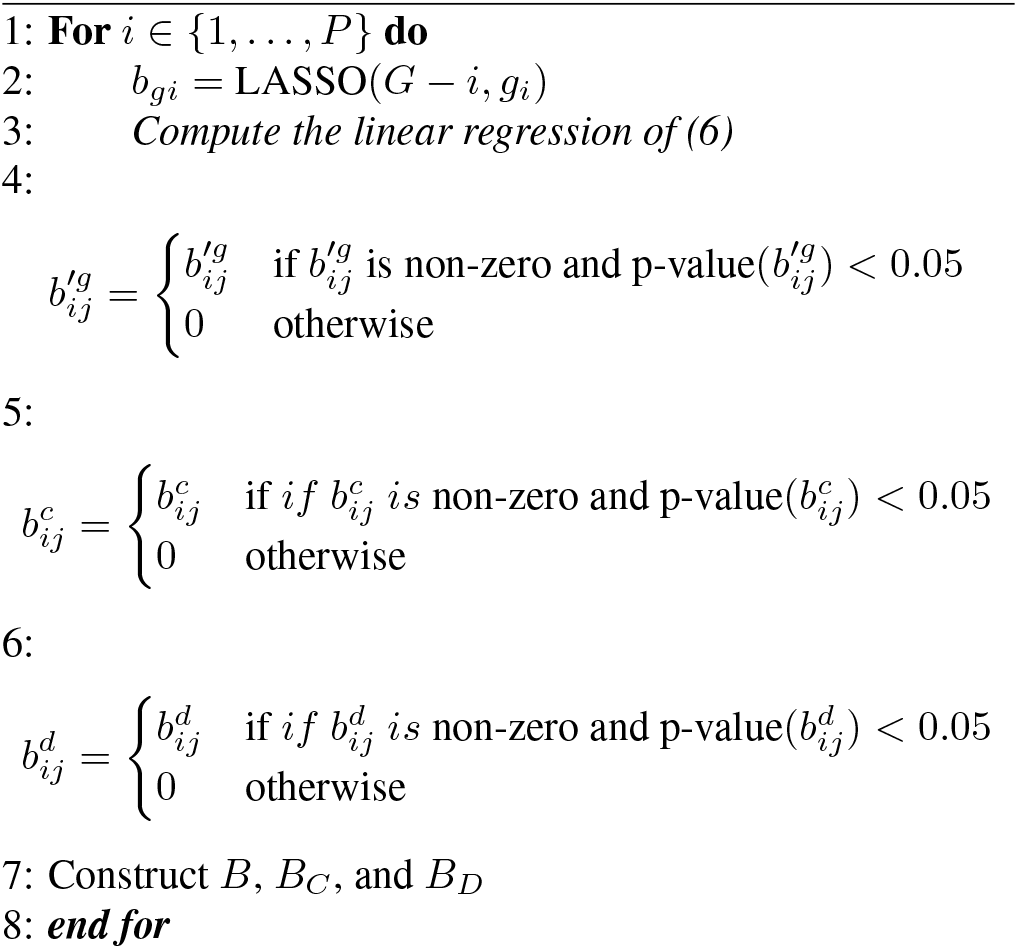

## 4. Simulation studies

We conducted intensive simulation experiments to evaluate our proposed method and compare the performance with existing methods. Due to only few available well-known true models of biological networks, the assessment of gene regulatory network inference in complex organisms such as human is challenging. Thus, the performance was indirectly evaluated with simulation data that implements integrative biological networks where the true model is given.

We generated the simulation data under the assumption that we hypothesised for the integrative gene regulatory networks. In the simulation studies, we aim to (1) verify that our proposed method produces robust performance to identify the true models of gene regulatory networks from multi-omics data, and (2) to compare the performance with current state-of-the-art methods on the given hypothesis. We carried out the following three experiments with the simulation data: (1) Receiver Operating Characteristic (ROC) curve, (2) sensitivity, and (3) false discovery rate.

### 4.1. Simulation settings

In the integrative gene regulatory network model, gene expression can be represented by two components: (1) gene expression regulated by other genes (*G*_*g*_) and (2) interactions of CNVs and DNA methylations (*G*_*i*_), as shown in (2):

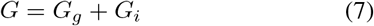

where

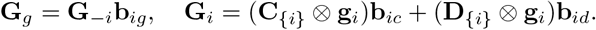

First, **G**_*g*_ was generated by the given adjacency matrix **Z**:

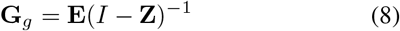

Where **I** ∈ ℝ^*N×P*^ is an identity matrix, and **E** ∈ ℝ^*N×P*^ is a matrix with normally distributed random values for noise, **E** ∼ *𝒩* (0, 0.01). The adjacency matrix **Z** is a sparse acyclic graph without self-loop.

The CNV data (**C** ∈ ℝ^*N×P*^) was implemented by taking the values {0, 1, 2, 3, 4} with the corresponding probabilities {0.01, 0.02, 0.4, 0.2, 0.1}. The given probabilities were directly acquired from CNV of human brain data. The DNA methylation (**D** ∈ ℝ^*N×M*^) was randomly obtained by the uniform distribution on the interval [0, 1]. In practice, CNVs and DNA methylations were annotated to nearby genes by using their loci and gene regions. We designated the associations by sparse Boolean mapping matrices **W** ∈ ℝ^*V ×P*^ and **F** ∈ ℝ^*M×P*^ for CNVs and DNA methylations, where only a couple of CNVs and DNA methylations can be annotated to a gene. In this simulation data, we assume that all of the CNVs and DNA methylations nearby a gene significantly regulate the gene expression.

The gene expression regulated by the interactions of CNVs and DNA methylations was generated by:

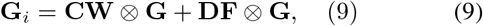

The gene expression controls the gene expression levels of other genes with the interaction effects of multi-omics data in gene regulatory networks. Therefore, we repeated Equation (8) and Equation (9) until **G** converges. Note that **Z, W**, and **F** are the (bi)adjacency matrices of ground truth in the simulation studies. The algorithm is described in Algorithm 2.

#### Algorithm 2 Algorithm 2

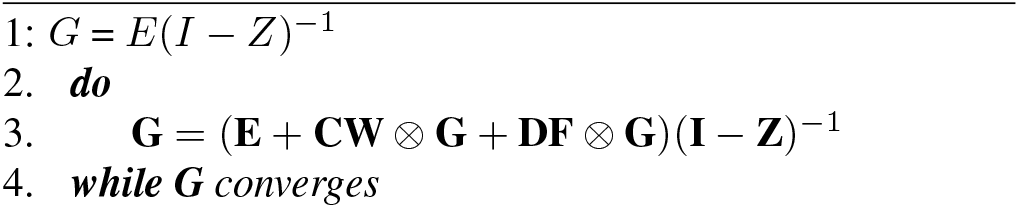

We considered a LASSO-based GRN method (GRN) as baseline and DCGRN (D.-C. Kim et al., 2014b) which is an integrative gene regulatory network inference method that uses multi-omics data. GRN infers the gene regulatory relationship on gene i with LASSO regularisation:

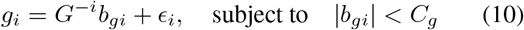

GRN identifies significant gene regulations by LASSO solution, but it considers only gene expression data for the network inference. In contrast, DCGRN incorporates multiomics data of CNVs and DNA methylations in the model:

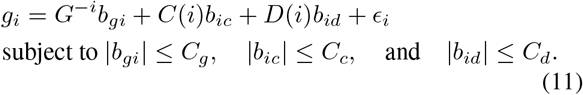

### 4.2. Experimental results with simulation data

First, we evaluated the performance by computing the area under the receiver operating characteristic curve (AUROC). The confusion matrix of true positive (TP), false positive (FP), true negative (TN), and false negative (FN) is defined as:

- TP: correctly identified the positive gene regulations as non-zero coefficients,
- FP: incorrectly identified the positive gene regulations as zero coefficients,
- TN: correctly identified the negative gene regulations as zero coefficients,
- FN: incorrectly identified the negative gene regulations as non-zero coefficients.

The non-zero coefficients of *b*_*gi*_, *b*_*di*_, and *b*_*ci*_ were considered as positives, while the coefficients of zero were negatives. The confusion matrices for gene regulations and integrative interactions of CNVs and DNA methylations were separately computed.

The ROC curves were traced over different thresholds to examine the trade-off between True Positive Rate (TPR = TP/(TP+FN)) and False Positive Rate (FPR = FP/(FP+TN)). The hyper-parameters (*λ*_*g*_, *λ*_*c*_, and *λ*_*d*_) in (3) determine the sparsity of significant components with non-zero coefficients in the multi-omics data. Note that all of the coefficients are non-zero when the parameter is zero, while all coefficient values become zero when an infinite value is given for the parameter. We considered the sparsity step (1 ≤ *θ* ≤ *P* + *V* + *M*) that determines the hyper-parameters in the LASSO solution. In this simulation study for the ROC curves, only the coefficient values were considered to determine the positive interactions, where p-values were not computed.

GRN computes only a confusion matrix for gene regulations, while DCGRN and MCNET have confusion matrices for CNVs and DNA methylations as well as gene expression. Therefore, overall ROC curves were considered, where only the confusion matrix of gene regulation was reflected on GRN, while the three confusion matrices were combined to compute ROC curves in DCGRN and MCNET. The overall ROC curves are illustrated in Figure 2, and AUROC is shown in Table 1. The experimental result of the overall AUROC supports that MCNET (0.938) provides better performance than GRN (0.895) and DCGRN (0.843).

**Table 1.**
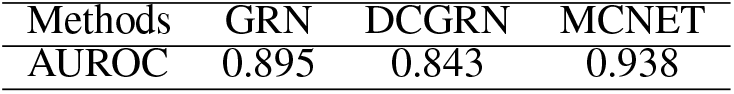
AUROC with simulation data

| Methods | GRN | DCGRN | MCNET |
| --- | --- | --- | --- |
| AUROC | 0.895 | 0.843 | 0.938 |

**Figure 2.**
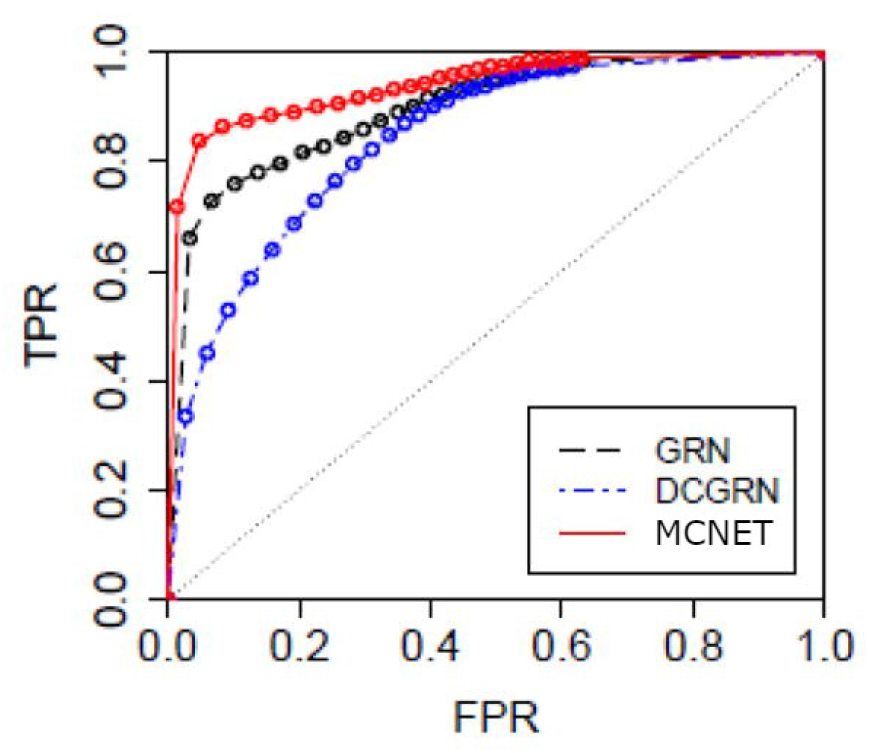
Overall ROC curves

TPRs on interactions of the CNVs and DNA methylations were measured for DCGRN and MCNET. Since the simulation data does not include negatives on CNVs and DNA methylations, we examined how well the methods identify the true positives. The TPRs are shown in Figure 3, where MCNET outperforms DCGRN in identifying true integrative interactions of CNVs and DNA methylations.

**Figure 3.**
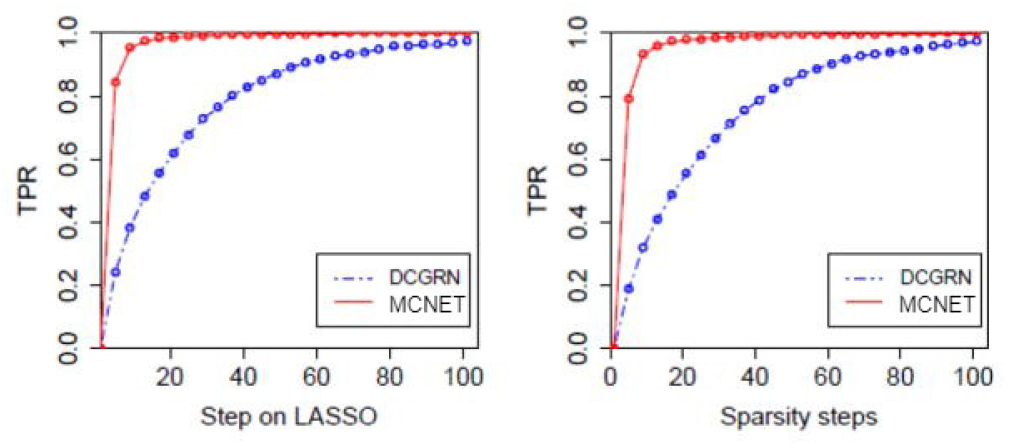
TPRs for interaction effects of CNVs and DNA methylations

Secondly, we measured the overall sensitivity which is the probability of identifying the true positives. In this simulation study, the hyper-parameters were optimised by 10-fold cross-validation. The multi-omics elements with non-zero coefficients and whose p-values are less than 0.05 are considered as positives. The overall sensitivity is depicted in Figure 4. MCNET produced the best sensitivity (0.300 ± 0.034), while GRN and DCGRN showed 0.199 ± 0.030 and 0.269 ± 0.035 respectively. The sensitivities of CNVs and DNA methylations on MCNET and DCGRN are shown in Figure 5. The sensitivities for MCNET and DCGRN were 0.102 ± 0.035 and 0.054 ± 0.030, respectively.

**Figure 4.**
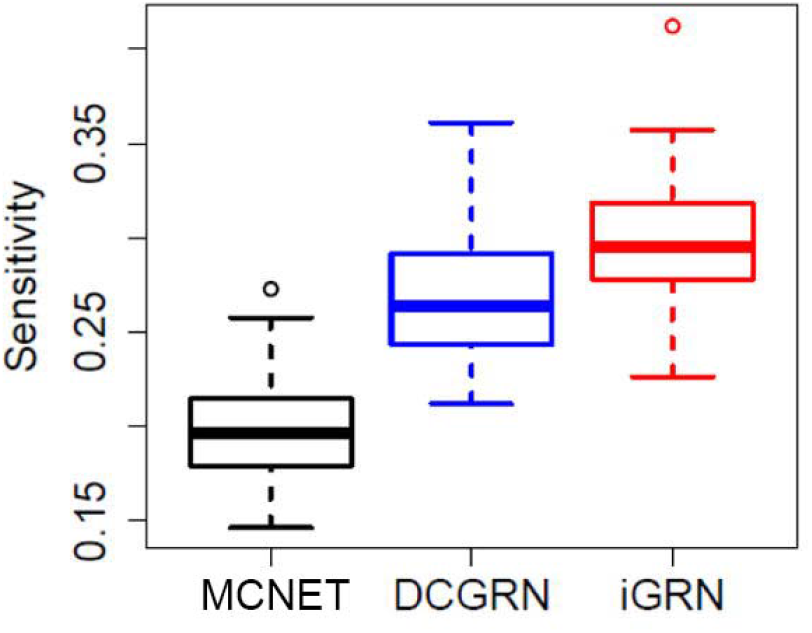
Sensitivity

**Figure 5.**
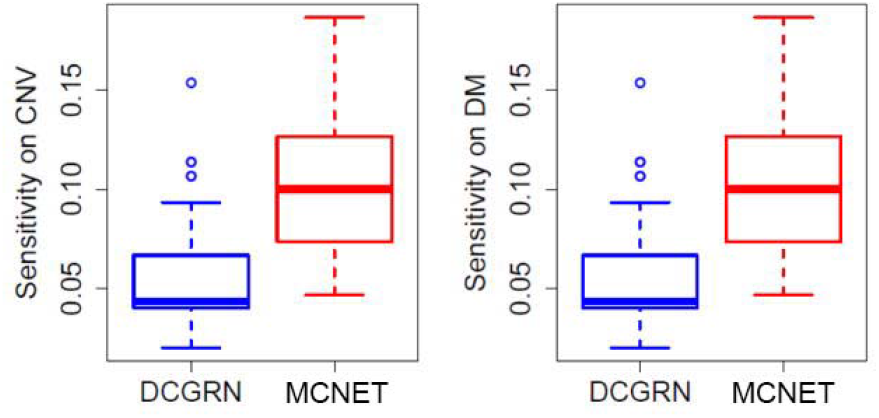
Sensitivity on copy number variations and DNA methylations

Lastly, we conducted the simulation study for False Discovery Rate (FDR). In this study, we generated simulation data that had no gene-gene regulation in the biological network. All positive predictions inferred by the methods were false positives, as the true adjacency matrix consisted entirely of zeros. FDR was computed as FP/(TP + FP). The FDRs of GRN, DCGRN, and MCNET, which were observed in Figure 6, were all less than 0.02. Specifically, the FDRs were 0.019 ± 0.003, 0.019 ± 0.003, and 0.019 ± 0.003 for GRN, DCGRN, and MCNET, respectively. These results indicate that MCNET had a chance of misidentifying interactions of less than 2

**Figure 6.**
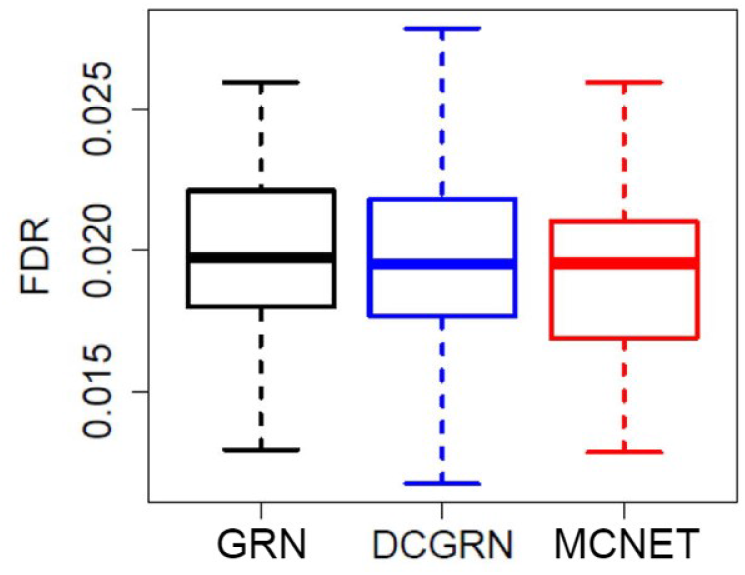
False discovery rate

## 5. Conclusion

The promise of deep learning, based on its success in other fields (Krizhevsky et al., 2017), is now also being seen across many different areas of biological network analysis. The methods we reviewed reported to consistently match or beat previous state-of-the-art methods using classical machine learning algorithms, providing evidence of one of deep learning’s core advantages: its strong empirical classification performance. Another advantage of deep learning is its ability to effectively deal with large datasets, which can be challenging for classical machine learning methods (Zhou et al., 2017) Although the training process of deep learning models with huge amounts of data is a non-trivial task, the advances in parallel and distributed computing have made training these large deep learning models possible (Dean et al., 2012; LeCun et al., 2015). The large number of matrix multiplications, high memory requirements and easy parallelizability of neural networks have been particularly well served by the recent breakthroughs in GPU computing (**?**). Finally, given that deep learning is a learning approach based on a hierarchy of non-linear functions, it is capable of detecting patterns in the raw data without explicit feature engineering. While it is not the only method that can handle non-linear relationships, the composition of many simple, non-linear layers makes it particularly adept at learning patterns at different layers of abstraction (LeCun et al., 2015), enabling more complex patterns to be detected. While deep learning methods are very promising, there are limitations and many open questions to be solved. One of the main problems with deep learning is its lack of interpretability. While there has been some recent progress in this area (Ching et al., 2018; Li et al., 2019), the black box nature of deep learning algorithms remains a key challenge, particularly in bioinformatics, where one is interested in understanding the mechanisms underlying the biological processes (Miotto et al., 2018; Zampieri et al., 2019). Additionally, interpretability is critical in the context of models that guide medical decisions, where doctors and patients are often unlikely to trust the output of a deep learning model without sufficient understanding of the prediction process (Ching et al., 2018).

Multi-omics data can be used in modeling gene regulatory networks. The recent rapid advances of high-throughput omics technologies have triggered the integrative multi-omics study for the in-depth understanding of the complex biological processes. However, only a few studies have considered the multi-omics data in gene regulatory network inference.

In this paper, we proposed an integrative gene regulatory network inference method, where multi-omics data and their interaction effects are integrated in the mathematical graph model. Our proposed method, MCNET, can infer gene regulatory networks from multi-omics data of CNVs and DNA methylations as well as gene expression data, and produce the homogeneous network where nodes are only genes. It enables one to analyse the gene regulatory network with most network analysis and visualisation tools efficiently. The inference capability of MCNET was assessed by the intensive experiments with simulation data. MCNET was applied to human brain data of psychiatric disorders, and the biological network of psychiatric disorders was analysed.

## Data Availability

All data produced in the present study are available upon reasonable request to the authors.

https://github.com/ansschh/MCNET.git

